# Improved characterization of circulating tumor cells and cancer-associated fibroblasts in breast cancer patients using imaging flow cytometry

**DOI:** 10.1101/2023.04.27.23289190

**Authors:** Anna Muchlińska, Robert Wenta, Wiktoria Ścińska, Aleksandra Markiewicz, Grażyna Suchodolska, Elżbieta Senkus, Anna J Żaczek, Natalia Bednarz-Knoll

## Abstract

Circulating tumor cells (CTCs) and circulating cancer-associated fibroblasts (cCAFs) have been individually considered as strong indicators of cancer progression. However, technical limitations have prevented their simultaneous analysis in the context of CTC phenotypes different from epithelial. This study aimed to analyze CTCs and cCAFs simultaneously in peripheral blood of 210 breast cancer patients using DAPI/pan-keratin (K)/vimentin (V)/alpha-SMA/CD29/CD45/CD31 immunofluorescent staining and novel technology - imaging flow cytometry (imFC). Single and clustered CTCs of different sizes and phenotypes (i.e. epithelial phenotype K+/V-, and epithelial-mesenchymal transition (EMT)-related such as K+/V+, K-/V+ and K-/V-) were detected in 27.6% of the samples and correlated with metastases. EMT-related CTCs interacted more frequently with normal cells and tended to occur in patients with tumors progressing during therapy, while cCAFs coincided with CTCs (mainly K+/V- and K-/V-) in 7 (3.3%) patients and seemed to correlate with the presence of metastases, particularly visceral ones. This study emphasizes advantages of imFC in the field of liquid biopsy and highlights the importance of multimarker detailed analysis of different subpopulations and phenotypes of cancer progression-related cells i.e. CTCs and cCAFs. Co-detection of CTCs and cCAFs might improve the identification of patients at higher risk of progression and their monitoring during therapy.

**Simple Summary:** Liquid biopsy is promising but challenging tool potentially upgrading cancer patients diagnostics and bringing new insights into tumor biology. Here, we applied a unique approach to detect CTCs and cCAFs in one-tube assay using imaging flow cytometry enabling improved enumeration, multimarker-based phenotyping and detailed morhopological characterization of those rare cells. We showed that EMT-related CTCs might contribute to breast cancer progression, whereas coincidence of CTCs and cCAFs might be signature of metastasis.

## Introduction

For last two decades liquid biopsy has been developed dynamically to provide solutions (i.e. ultrasensitive technologies and/or specific markers) relevant for early diagnostics and monitoring of cancer patients ^1^. Circulating tumor cells (CTCs), though very plastic and heterogenous due to e.g. epithelial-mesenchymal transition (EMT) ^2, 3^, have been shown to be a non-invasive, prognostic biomarker in many solid tumors, including breast cancer (BC), one of the most common cancers worldwide among female population. Yet, other components released from primary or secondary tumors, such as circulating endothelial cells, cancer-associated macrophage-like cells or fibroblasts (CAFs), as well as extracellular vesicles can also be found in blood of cancer patients and putatively correlate with worse prognosis ^4–6^.

Key players in dissemination and, more importantly, in metastasis formation are naturally CTCs, the “seeds” of new lesions ^7^. In concordance, the poor prognostic connotation of CTCs was demonstrated in both early ^8^ and metastatic breast cancer ^9^ and many other solid tumors ^10–12^. In particular, intermediate and full EMT-related phenotypes (i.e. epithelial-mesenchymal and mesenchymal phenotype) have been identified as potentially more aggressive ones, and possibly even more importantly - difficult to detect using common epithelial markers-dependent methods ^2, 13^. Furthermore, it was shown in a mouse model of lung cancer metastases that CTCs can also carry stromal cells, including activated fibroblasts, as fragments of "soil" ready to promote metastasis formation ^14^. In accordance with this model, cCAFs were observed in blood of cancer patients both individually and clustered with CTCs ^6, 15, 16^. Interestingly, cCAFs were isolated mostly from blood samples of metastatic cancer patients, whereas CTCs were detected in blood samples of patients at all stages of disease, including early cancers ^6, 15, 17^.

Of note, co-detection of CTCs and cCAFs in one-tube assay has been difficult as multimarker-based identification of rare cells in blood is still limited, even using golden standard CellSeach. The exact co-detection of CTCs and cCAFs was restricted exclusively to epithelial CTCs, missing information about potential EMT-related phenotypes of CTCs ^6, 16^. Meanwhile, new technologies, not relying on exclusively epithelial markers, such as imaging flow cytometry (imFC) ^18^ might enable detailed analysis of different subpopulations of cells in one-tube assay in relatively short time. As imFC offers combination of the features of classical flow cytometry (including an impartial multiparameter and high-throughput analysis) and high resolution fluorescence microscopy (allowing insight into morphology of each individual examined object), it might boost investigation on tumor dissemination.

Therefore, in the present study, the feasibility of imFC for simultaneous CTC and cCAF detection from blood samples of breast cancer patients has been evaluated. Furthermore, clinical relevance of those rarely detected cells has been assessed.

## Methods

### Breast cancer patients and non-cancer volunteers cohort

Two-hundred-ten female breast cancer patients (age ≥ 18, with no coexisting malignancy) treated in the Breast Unit, University Clinical Center in Gdańsk between 2019-2022 and 20 female volunteers (age ≥ 18) with no cancer history (further named as healthy volunteers) from Central Clinical Laboratory, University Clinical Center in Gdańsk were recruited for this study after providing written informed consent. Clinico-pathological parameters, pretreatment blood count (incl. leukocytes and their subtypes) and response to therapy were documented for the breast cancer patients **(Table 1)**. The study was conducted in accordance with the Declaration of Helsinki and approved by the Independent Bioethics Committee for Scientific Research at Medical University of Gdańsk (protocol no. NKBBN/748/2019-2020).

**Table 1.**
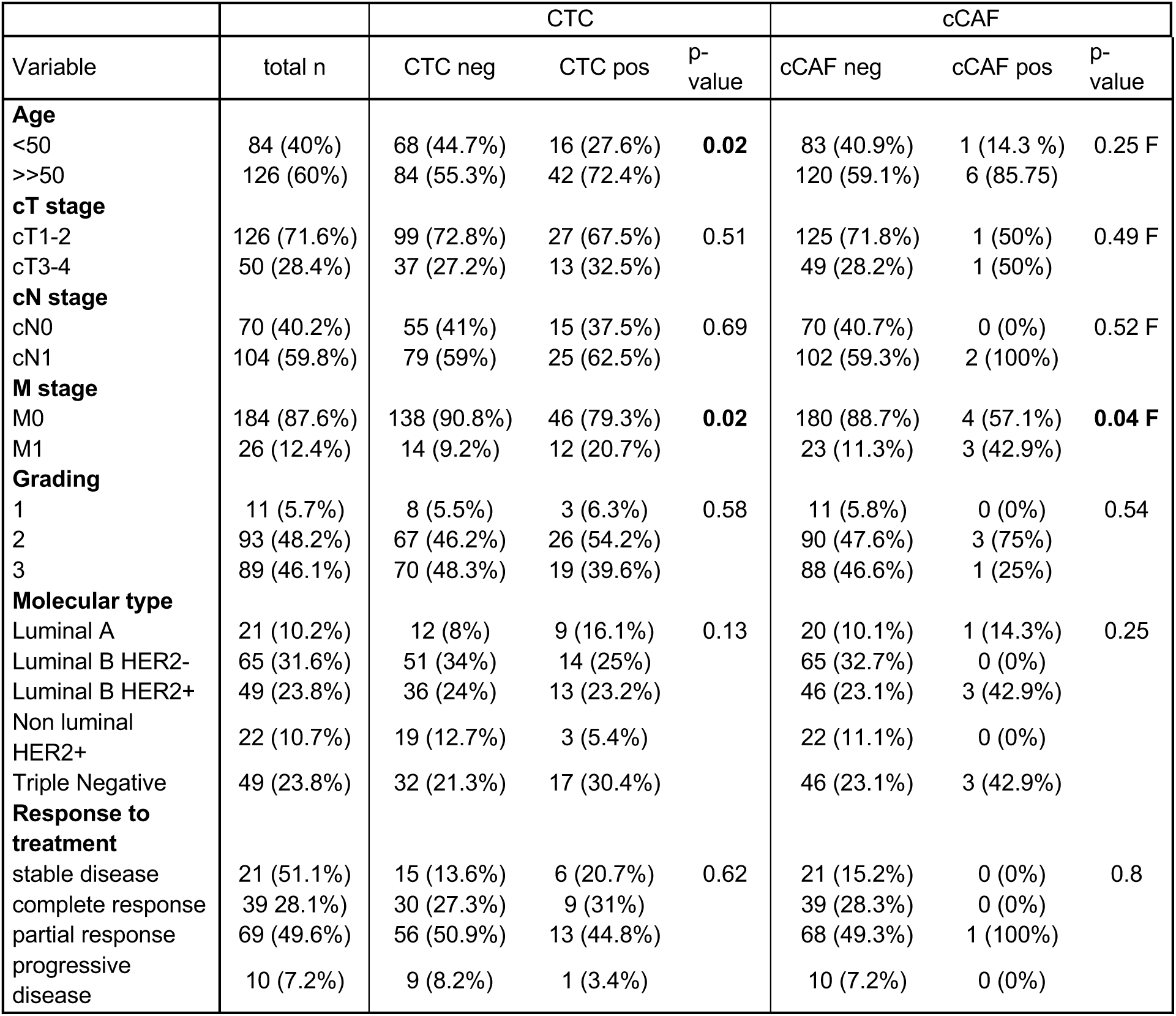
Comparison of CTCs and cCAFs status to clinico-pathological features and response to treatment in breast cancer patients. F indicates Fisher’s exact test, otherwise Chi squared test was performed. Due to missing data not all numbers sum up to 210.

| Variable | total n | CTC |  |  | cCAF |  |  |
| --- | --- | --- | --- | --- | --- | --- | --- |
|  |  | CTC neg | CTC pos | p-value | cCAF neg | cCAF pos | p-value |
| **Age** |  |  |  |  |  |  |  |
| <50 | 84 (40%) | 68 (44.7%) | 16 (27.6%) | **0.02** | 83 (40.9%) | 1 (14.3 %) | 0.25 F |
| >>50 | 126 (60%) | 84 (55.3%) | 42 (72.4%) |  | 120 (59.1%) | 6 (85.75) |  |
| **cT stage** |  |  |  |  |  |  |  |
| cT1-2 | 126 (71.6%) | 99 (72.8%) | 27 (67.5%) | 0.51 | 125 (71.8%) | 1 (50%) | 0.49 F |
| cT3-4 | 50 (28.4%) | 37 (27.2%) | 13 (32.5%) |  | 49 (28.2%) | 1 (50%) |  |
| **cN stage** |  |  |  |  |  |  |  |
| cN0 | 70 (40.2%) | 55 (41%) | 15 (37.5%) | 0.69 | 70 (40.7%) | 0 (0%) | 0.52 F |
| cN1 | 104 (59.8%) | 79 (59%) | 25 (62.5%) |  | 102 (59.3%) | 2 (100%) |  |
| **M stage** |  |  |  |  |  |  |  |
| M0 | 184 (87.6%) | 138 (90.8%) | 46 (79.3%) | **0.02** | 180 (88.7%) | 4 (57.1%) | **0.04 F** |
| M1 | 26 (12.4%) | 14 (9.2%) | 12 (20.7%) |  | 23 (11.3%) | 3 (42.9%) |  |
| **Grading** |  |  |  |  |  |  |  |
| 1 | 11 (5.7%) | 8 (5.5%) | 3 (6.3%) | 0.58 | 11 (5.8%) | 0 (0%) | 0.54 |
| 2 | 93 (48.2%) | 67 (46.2%) | 26 (54.2%) |  | 90 (47.6%) | 3 (75%) |  |
| 3 | 89 (46.1%) | 70 (48.3%) | 19 (39.6%) |  | 88 (46.6%) | 1 (25%) |  |
| **Molecular type** |  |  |  |  |  |  |  |
| Luminal A | 21 (10.2%) | 12 (8%) | 9 (16.1%) | 0.13 | 20 (10.1%) | 1 (14.3%) | 0.25 |
| Luminal B HER2- | 65 (31.6%) | 51 (34%) | 14 (25%) |  | 65 (32.7%) | 0 (0%) |  |
| Luminal B HER2+ | 49 (23.8%) | 36 (24%) | 13 (23.2%) |  | 46 (23.1%) | 3 (42.9%) |  |
| Non luminal HER2+ | 22 (10.7%) | 19 (12.7%) | 3 (5.4%) |  | 22 (11.1%) | 0 (0%) |  |
| Triple Negative | 49 (23.8%) | 32 (21.3%) | 17 (30.4%) |  | 46 (23.1%) | 3 (42.9%) |  |
| **Response to treatment** |  |  |  |  |  |  |  |
| stable disease | 21 (51.1%) | 15 (13.6%) | 6 (20.7%) | 0.62 | 21 (15.2%) | 0 (0%) | 0.8 |
| complete response | 39 (28.1%) | 30 (27.3%) | 9 (31%) |  | 39 (28.3%) | 0 (0%) |  |
| partial response | 69 (49.6%) | 56 (50.9%) | 13 (44.8%) |  | 68 (49.3%) | 1 (100%) |  |
| progressive disease | 10 (7.2%) | 9 (8.2%) | 1 (3.4%) |  | 10 (7.2%) | 0 (0%) |  |

### Blood collection and processing for CTCs enrichment

EDTA-peripheral blood samples (vol. of 7.5 ml) were collected from all recruited patients and processed as soon as possible after blood donation (with 68% of samples processed within up to 3 hours after donation). First 3 ml of blood were discarded to avoid skin cells (i.e. keratinocytes, fibroblasts) and endothelial cells contamination during the puncture. Peripheral blood mononuclear cells (PBMCs) fraction was isolated using density gradient centrifugation. Briefly, blood was centrifuged at 200g for 10 min at room temperature (RT) to separate platelets-rich-plasma. Then, remaining of the blood sample was diluted with 1x phosphate buffered saline (1xPBS) up to 9 ml, layered onto the Histopaque®-1077 (Sigma-Aldrich) and centrifuged at 400g for 30 min with break off. PBMCs fraction was harvested, fixed in 4% formaldehyde and stored at -80°C in 0.5ml aliquots for further analysis.

### Cell culture

MCF7 (HTB-22), T-47D (HTB-133), BT-474 (HTB-20), MDA-MB-361 (HTB-27), SKBR3 (HTB-30), HCC1806 (CRL-2335), MDA-MB-231 (HTB-26) cells were purchased from the American Tissue Culture Collection (ATCC) and used for immunofluorescent staining protocol optimization. Cells were cultured in Dulbecco’s Modified Eagle Medium (DMEM) supplemented with 10% fetal bovine serum (FBS) under appropriate conditions and routinely tested for mycoplasma contamination. For all cell lines, the same batch of FBS was used. Cells were cultured until 80% of confluence, trypsynized, fixed in 4% formaldehyde and stored at -80°C for further analysis.

### Immunofluorescent staining for simultaneous CTCs and cCAFs detection

Immunofluorescent staining of different breast cancer cell lines and clinical samples was performed using cocktail of antibodies aiming to detect CTC markers: pan-keratins (K, AE1/AE3 clone, AF488-conjugated, Thermo Fisher Scientific, #53-9003-82 + C11 clone, AF488-conjugated, Thermo Fisher Scientific, #MA5-18156) and vimentin (V, D21H3 clone, AF647-conjugated, Cell Signalling, #9856); cCAF markers: alpha smooth muscle actin (α-SMA, 1A4 clone, PE-conjugated, R&Dsystems, #IC1420P) and CD29 (TS2/16 clone, SuperBright600-conjugated, Thermo Fisher Scientific, #63-0299-42); leukocyte marker: CD45 (REA747 clone, APC-Vio770-conjugated, Miltenyi Biotec, #130-110-635); as well as endothelial cell marker: CD31 (WM59 clone, APC-Cy7-conjugated, BioLegend, #303120) diluted 1:2500, 1:2500, 1:50, 1:100, 1:1000, 1:50, 1:10, respectively (prepared in 1x Perm-Wash Buffer, BD Biosciences). Investigated cells were thawed and washed with 1ml of 1x PBS to remove formaldehyde, and then incubated for 30 min at +4°C with antibody cocktail. After one additional washing step in 1x PBS, cells were resuspended in 30 μl of 1x PBS, counterstained with DAPI (BD Biosciences, 1μg/ml) and immediately analyzed using Amnis® ImageStream®X Mk II (Luminex).

### Imaging flow cytometry analysis

Amnis^®^ ImageStream^®^ X Mk II (Luminex), equipped with lasers at 405 nm, 488 nm and 642 nm and the INSPIRE™ software (version 200.1.681.0; Luminex) was used for samples analysis and data acquisition. Subsequent analysis of the obtained outcomes was performed with the IDEAS software (version 6.2; Luminex). Cells were imaged at 40x magnification at low speed for receiving high–quality images. The optimal compensation matrix between individual fluorescence channels was established using a mixture of MDA–MB–231 cells, MCF7 cells and PBMCs from a healthy donor, stained with V, CD45, CD31 antibodies and DAPI separately or in combination. The optimized parameters setting for acquisition was used for all analyzed samples (**Supplemental Table 1**).

All DAPI+ cells were counted and considered as the initial gate, whereas DAPI+CD45&CD31-cells were collected. To gate potential CTCs, fluorescence intensity of K and V were visualized in a 2D dot plot. To gate cCAFs, fluorescence intensity of α-SMA and CD29 was visualized in histograms and cut at 10^4^. Potential preselected CTCs and cCAFs were visually confirmed from immunofluorescence images for their morphology and staining details, tagged, and counted manually.

Detected cells were defined as: epithelial CTCs if K+/DAPI+/V-/CD45&CD31-(abbreviated to K+V-), mesenchymal CTCs - K-/DAPI+/V+/CD45&CD31- (K-V+), epithelial-mesenchymal CTCs - K+/DAPI+/V+/CD45&CD31- (K+V+), negative for both epithelial and mesenchymal markers - K-/DAPI+/V-/CD45&CD31-(K-V-), as well as cCAFs - α-SMA+/K-/DAPI+/V- or V+/CD45&CD31- or CD29+/K-/DAPI+/V- or V+/CD45&CD31-phenotype was detected. CTCs dimensions such as diameter, circumference, area, and circularity were measured using QuPath ver. 0.2.3 software ^19^. Cytoplasmatic protrusions were defined as fragments of cell membrane extending beyond the estimated overall cell geometry.

Presence, numbers, clusters, and phenotypes of CTCs or cCAFs as well as their morphological details or interactions were further correlated with clinico-pathological data (i.e. age, T, N and M status, grading, hormone receptor (HR) and HER2 status, molecular subtype, pretreatment blood count and response to therapy).

### Statistics

Statistical analysis was performed with SPSS 27.0.1.0 software package (SPSS Inc., Chicago, IL, USA) licensed for the University of Gdańsk. The exact numbers of detected CTCs and cCAFs were calculated per 1 million of PBMCs to normalize all samples. Different classifications of patients in regard to their CTCs status were investigated, including exclusive CTCs phenotype, dominant CTCs phenotype and epithelial vs. EMT-related phenotype assigned to an individual patient. Chi squared, Fisher’s exact or Fisher-Freeman-Haltman tests were used to compare CTCs and/or cCAFs-positive or -negative patients and patients with different CTC phenotypes with clinico-pathological parameters (age, T stage, N stage, M stage, grade, molecular subtype, and response to treatment). Differences in enumeration of CTCs and cCAFs between BC patients group with different clinico-pathological parameters were assessed by Mann-Whitney or Kruskal-Wallis tests. All statistical analyses were two-sided, and p<0.05 was designated as statistically significant.

## Results

### Detection and characterization of different phenotypes of circulating tumor cells and circulating cancer-associated fibroblasts

CTCs were not detected in 20 samples of female heathy volunteers (age range 18-77, median 40), whereas breast cancer cell lines of different molecular subtypes revealed expected epithelial or EMT-related phenotypes (**Fig. S1**).

All samples from 210 breast cancer patients were informative. One-hundred-eighty-three patients were analyzed prior to therapy, 24 were before the subsequent line of therapy, whereas for 3 patients those data were missing. Cells isolated from blood were initially gated as DAPI+ and CD45/CD31- and different sorts of investigated cells were identified within this population. Putative CTCs were detected in 58 (27.6%) of 210 blood samples. Their numbers ranged between 1−459/1 mln of PBMCs (median 10/1 mln of PBMCs). Among those putative CTCs 4 phenotypes were identified (**Fig. 1A**): epithelial CTCs (i.e. K+/V-), and EMT-related CTCs such as epithelial-mesenchymal (epi/mes) CTCs (i.e. K+/V+), mesenchymal CTCs (i.e. K-/V+), and negative for both epithelial and mesenchymal markers (i.e. K-V-; named further as negative) CTCs. They occurred in 30 (14.3%), 26 (12.4%), 12 (5.7%), 21 (10%) of 210 BC patients, respectively. Putative epithelial-mesenchymal and mesenchymal CTCs seemed to be smaller than epithelial CTCs, whereas negative CTCs were their size (**Fig. 1B**). Epithelial-mesenchymal and negative CTCs revealed protrusions more frequently than epithelial CTCs or mesenchymal CTCs (**Fig. 1C**). None of CTCs expressed CD29 (data not shown).

**Figure 1.**
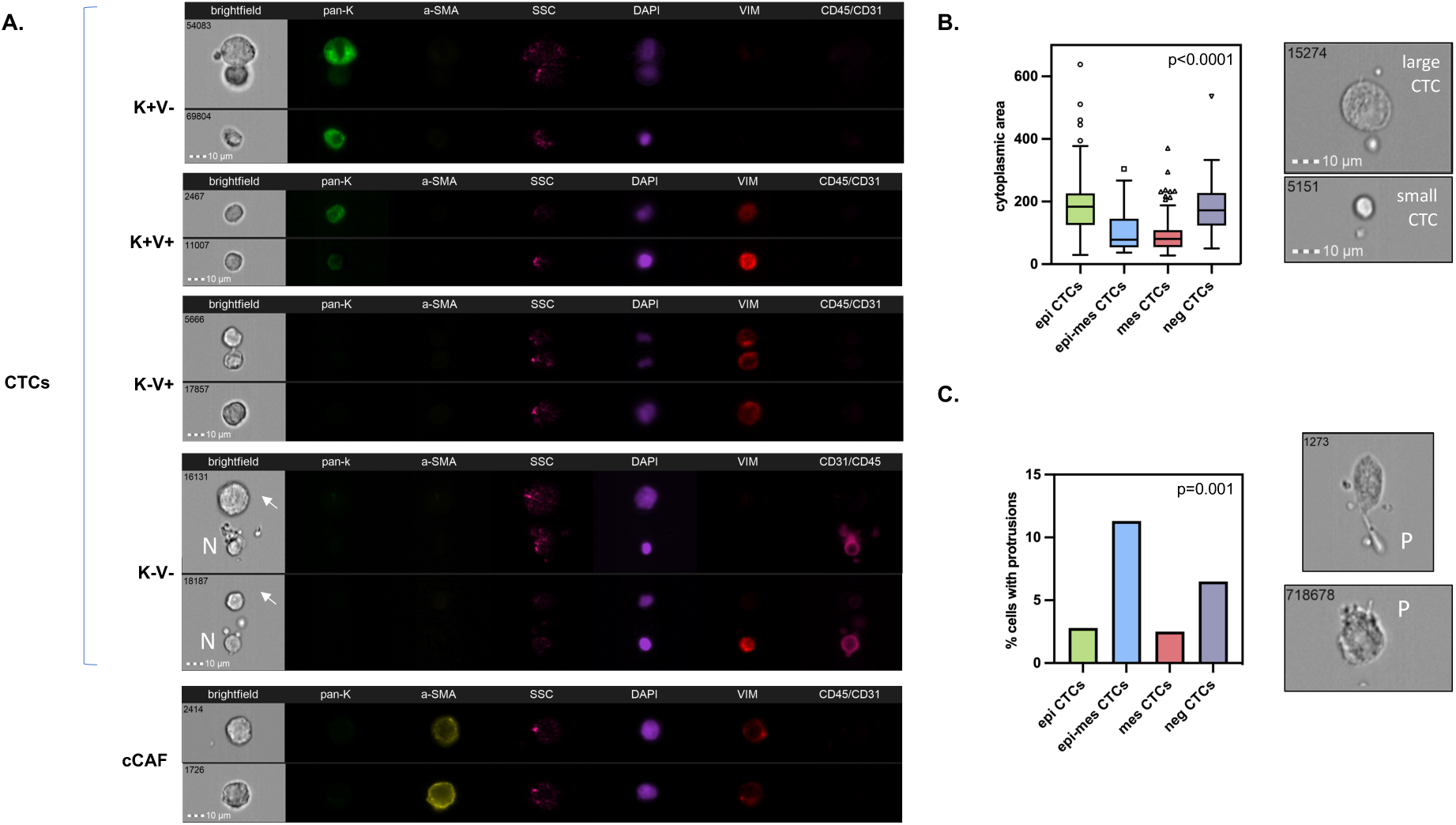
Characterization of CTCs in breast cancer: examples of their different phenotypes (**A**), comparison of their sizes defined as cytoplasmic area (**B**) and occurrence of protrusions among them (**C**). BF indicates brightfield, pan-K – pan-keratins, a-SMA – alpha-smooth muscle actin, SSC – side scatter, Vim – vimentin, CD45/CD31 – leukocyte / endothelial cell marker, N – normal cell, arrows – CTC; all images are captured with objective 40x.

In addition, cCAFs were determined simultaneously in the same samples (**Fig. 1A**). cCAFs were not detected in 20 healthy donors. cCAFs classified as α-SMA+/K-/DAPI+/V- or V+/CD45&CD31- were present only in 7 (3.3%) of 210 breast cancer patients (range 7–56 cCAFs/1 mln of PBMCs and median 25 cCAFs/1 mln of PBMCs). They were characterized as V+ (13%) and V- (87%) and none of detected cCAFs expressed CD29. No CD29+/α-SMA-/K-/DAPI+/V- or V+/CD45&CD31-cCAFs were observed. They coincided with CTCs, in particular epithelial and negative CTCs (R^2^=0.165, p=0.017 and R^2^=0.250 = p<0.001, respectively, **Supplemental Table 2**). They were observed also more frequently in blood samples with the higher yields of CTCs (R^2^=0.198, p=0.004).

Of note, the numbers of identified CTC phenotypes and cCAFs were not associated with the time between blood sample donation and processing (range 18-1488 minutes, median 65 minutes), total number of PBMCs in the analyzed sample, neither counts of any leukocyte fractions performed in parallel during pretreatment blood count (data not shown).

Multiple clusters (range 2-11) of 2-3 CTCs occurred in 8 (3.81%) patients. They consisted mainly of solely i) epithelial, ii) epithelial-mesenchymal or iii) negative CTCs (**Fig. 2A-B**). Heterogenous CTC clusters of mesenchymal CTCs and negative CTCs were detected in one patient (**Fig. 2A-B**), and in two patients two homogenous CTC clusters of different phenotypes (epithelial and negative cells) were present. In addition, clusters of CTCs (only mesenchymal or negative ones) with normal cells were identified in 4 (1.90%) patients (**Fig. 2A-B**). The majority of those CTCs-normal cells clusters also contained platelets (**Fig. 2A-B**). In all of those 4 patients, clusters of CTCs and clusters of CTCs and normal cells were co-detected. No clusters of CTCs and cCAFs were detected.

**Figure 2.**
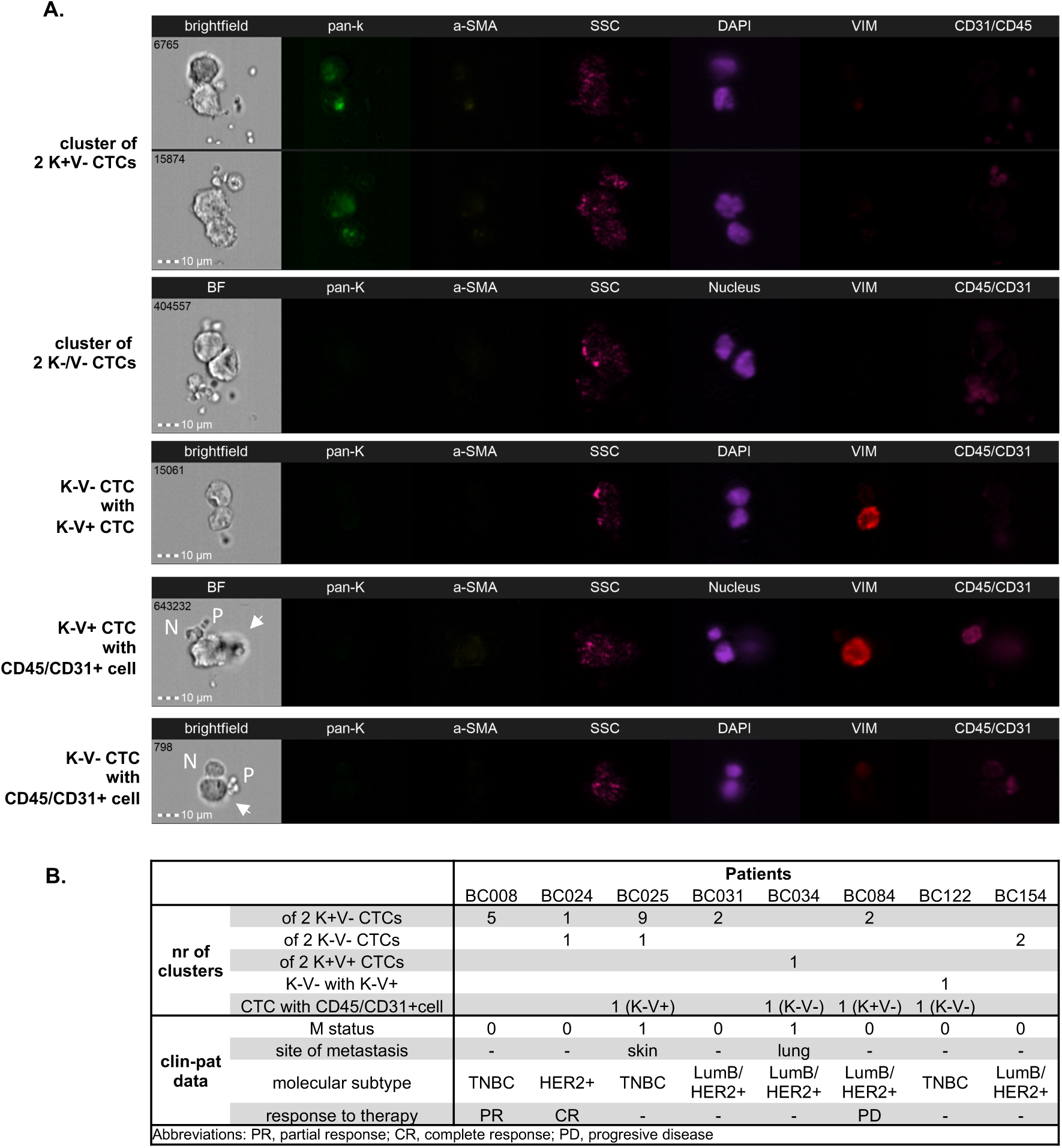
CTCs and CTCs-normal cells clusters in breast cancer detected using imaging flow cytometry: homogenous, heterogenous clusters, and CTCs clusters with normal cell (**A**). Clinico-patological data of clusters positive patients (**B**). BF indicates brightfield, pan-K – pan-keratins, a-SMA – alpha-smooth muscle actin, SSC – side scatter, Vim – vimentin, CD45/CD31 – leukocyte / endothelial cell marker, N – normal cell, P – platelets, arrows – CTC; all images are captured with objective 40x.

### Distribution of different phenotypes of circulating tumor cells and circulating cancer-associated fibroblasts

One CTC/1 mln of PBMCs was found in only 2 (1%) patients, whereas 56 (26.6%) patients had 2 or more CTCs/1 mln of PBMCs (including 36/17.1% patients with 5 or more CTCs/1 mln of PBMCs). Among the samples with 2≥CTCs, 33 (58.9% of CTCs-positive and 15.7% of all patients) and 23 (41.1% of CTCs-positive and 11% of all patients) seeded CTCs of one phenotype or different phenotype (i.e. homo- and heterotypic dissemination), respectively. Epithelial and mesenchymal phenotypes of CTCs were predominant, with epithelial-mesenchymal and negative CTCs occurring very rarely, and this effect was even more pronounced when the most enriched phenotype within the sample was considered in CTCs phenotype classification (**Fig. 3A-B**)

**Figure 3.**
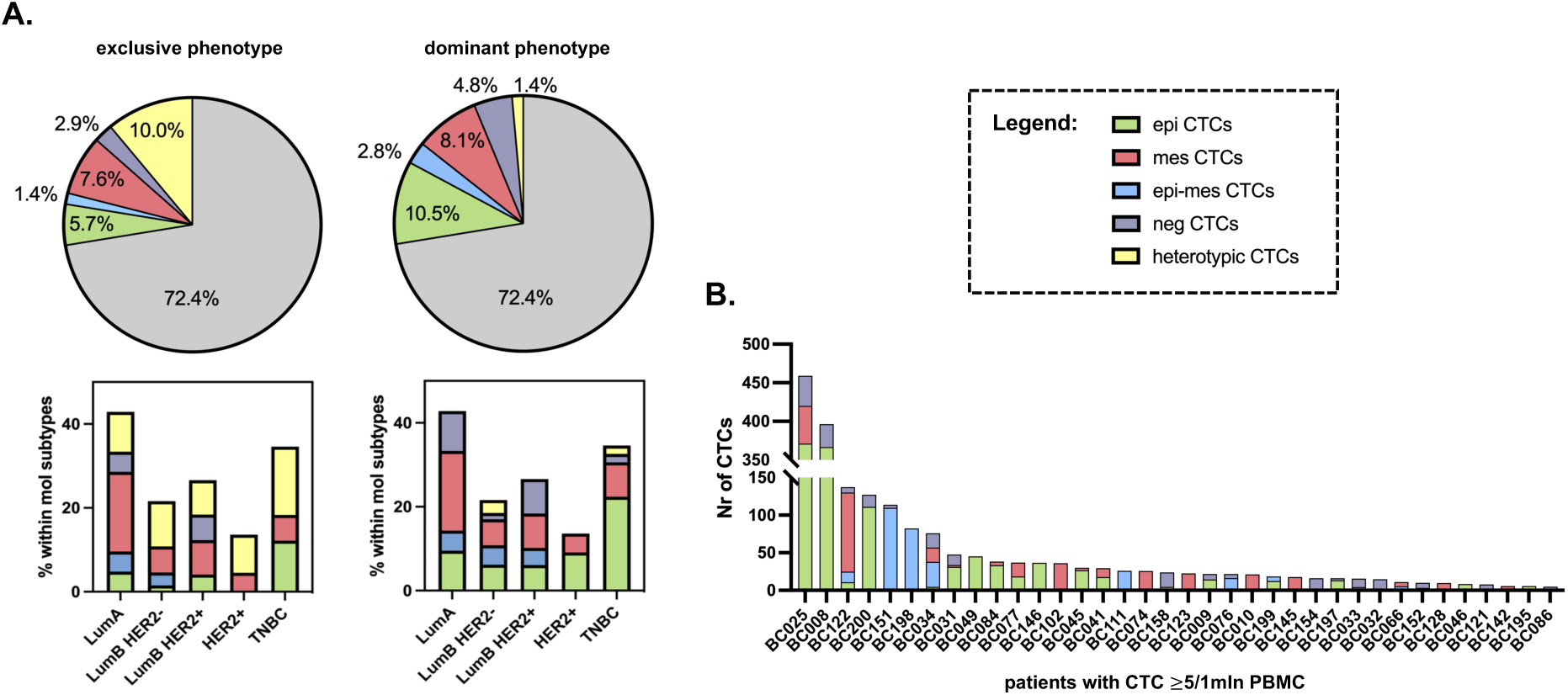
Distribution of CTCs phenotypes in breast cancer patients: distribution of exclusive and dominant CTCs phenotypes in the whole cohort of patients and among different molecular subtypes of breast cancer (**A**) and heterogeneity of CTCs in individual patients with CTC≥5 (**B**)

In total, when CTC-negative and -positive cases were considered, EMT-related CTCs occurred more frequently in luminal B/Her2+ (LumB/Her2+) BC subtype (20.4% of cases), whereas epithelial CTCs in TNBC (22.4% of cases) (Chi2 test, p=0.047, **Supplemental Table 3-4**). However, no statistically significant pattern of individual CTCs phenotype distribution was observed when exclusive or dominant phenotype classification was considered (**Fig. 3A**). Epithelial CTCs occurred more frequently (i.e. in 82% of 17 CTC-positive TNBC patients, p=0.021; data not shown) and at the higher numbers (Kruskal-Wallis test, p=0.013) in TNBC, whereas mesenchymal CTCs occurred almost equally frequently among all molecular subtypes (on average in 44.4% of CTC-positive patients).

cCAFs occurred mainly in LumB/Her2+ (n=3) and TNBC (n=3) patients. CTCs clusters occurred more frequently in samples with the higher yields of CTCs, heterotypic spread and presence of cCAFs (all p<0.001), independent of molecular subtype (data not shown).

### Clinical relevance of different phenotypes of circulating tumor cells and circulating cancer-associated fibroblasts

Presence, numbers, clusters, and phenotypes of CTCs or cCAFs were analyzed in relation to clinico-pathological parameters and patients’ response to therapy in the whole cohort of patients (**Table 1, Supplemental Table 3-4**) as well as in the subcohorts of patients defined based on the age of the patients, molecular subtype, N and T status, grade, as well as therapy timepoint (data not shown).

CTCs were detected more frequently in older patients (≥50 years old; p=0.023, **Table 1**). All phenotypes of CTCs were detected at all stages of tumor development (cT1-4) and lymph nodes involvement (cN0-3), whereas cCAFs (**Table 1**) and CTCs clusters were found in ≥cT2 tumors. Negative CTCs correlated with the higher number of involved lymph nodes (Fisher-Freeman-Haltman, p=0.048) (**Fig. 4A**).

**Figure 4.**
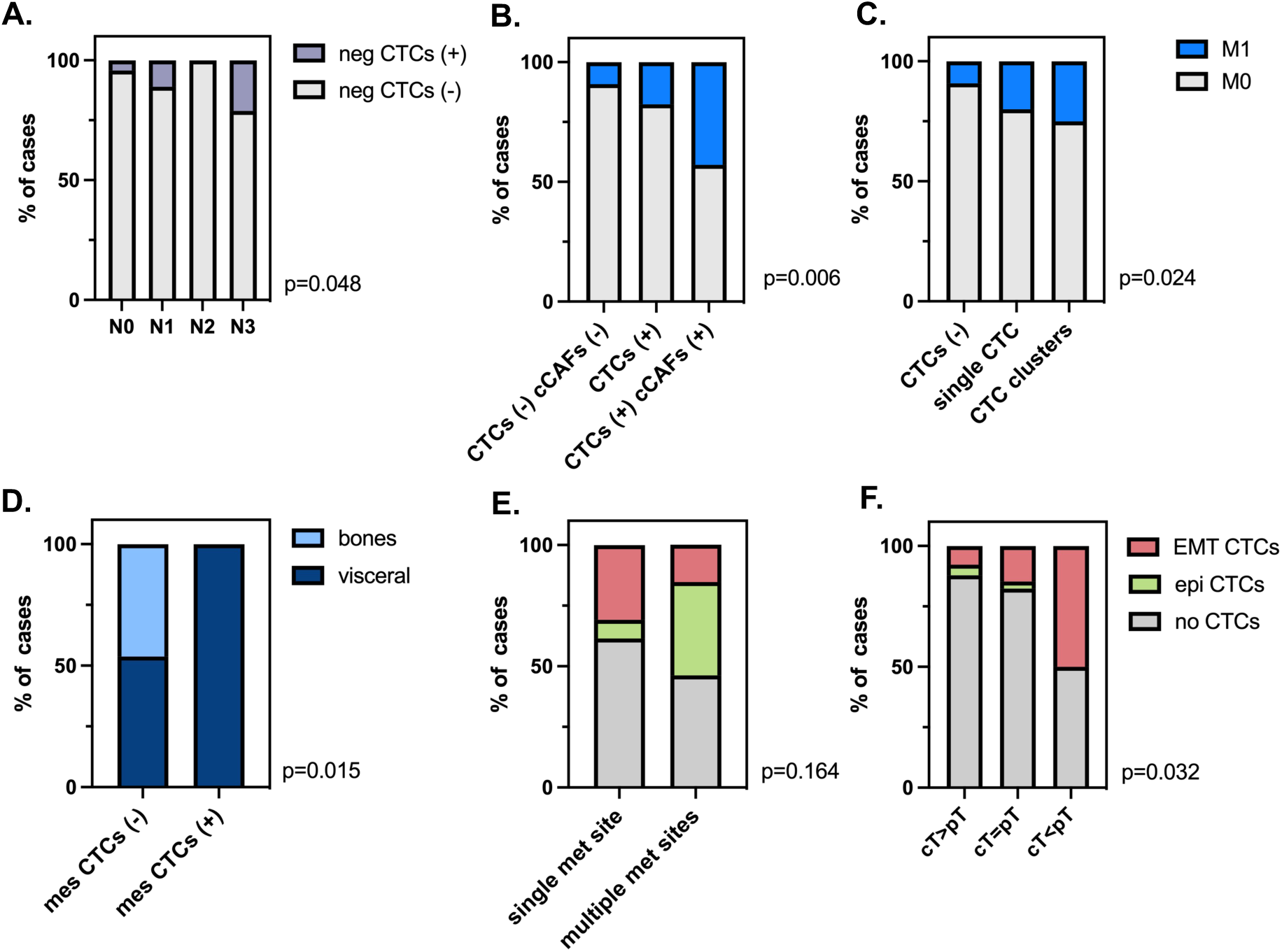
Clinical relevance of different phenotypes of CTCs and cCAFs: distribution of negative (neg) CTCs by N status **(A)**, presence of metastases depending on CTCs/cCAFs **(B)** and single/clustered CTC status **(C)**, mesenchymal (mes) CTCs correlation with a site of metastases **(D)** and CTCs phenotypes correlation with number of metastases **(E)** and with response to therapy **(F)**.

Both CTCs and cCAFs presence correlated with the presence of metastases (Chi^2^ test, p=0.024 and Fisher test, p=0.042, respectively) (**Table 1**). It seemed that particularly the presence of epithelial CTCs was associated with M1 status (p=0.033, data not shown). Frequency of metastases increased gradually between i) CTC-negative patients < CTC-positive patients < patients positive for both CTCs and cCAFs, as well as ii) CTC-negative patients < patients positive for single CTCs < patients positive for clustered CTCs, despite of the low number of cases in the last subgroups of patients in each classification (Fisher-Freeman-Haltman test, p=0.006 and p=0.024, respectively, **Fig. 4 B-C**). Of note, mesenchymal CTCs, cCAFs, and CTCs clusters were observed exclusively in patients with visceral metastases (but not of specific localization) (mesenchymal CTCs: Fisher’s exact test, p=0.015, **Fig. 4D;** cCAFs, and CTCs clusters: both p>0.05, data not shown), whereas epithelial CTCs – more frequently in patients with multiple metastatic sites (p>0.05, **Fig. 4E**), including solely visceral metastases or those coexisting with bone metastases (data not shown).

None of the investigated parameters (i.e. presence, numbers, clusters and phenotypes of CTCs, or cCAFs) correlated with the response to therapy either in the whole cohort of patients or only in patients with neoadjuvant (n=122) treatment. Nevertheless, EMT-related CTCs were the most enriched in the patients with tumors progressing during treatment (cT < pT, Chi^2^ test, p=0.032, **Fig. 4F**).

## Discussion

Rare cancer-related cells isolated from peripheral blood of cancer patients such as predominantly CTCs but also cCAFs are considered strong indicators of unfavorable prognosis. Here, they were investigated together in one-tube assay using novel method, imFC ^18^. This method warranted the simultaneous detection and detailed characterization of cCAFs and both epithelial and EMT-related phenotypes of CTCs overcoming technical limitations of current CTC technology.

CTCs were enumerated at frequency (approx. 25% and 46% of M0 and M1 BC patients, respectively) and yields (1−459/1 mln of PBMCs) similar to previously published data obtained using different methodology, including the FDA-approved “golden standard” of CTC technology, CellSearch ^20–23^. Of note, we identified four, rather than typically observed three, phenotypes ^24, 25^ related to the status of common epithelial (K) and mesenchymal (V) markers, including a K-V-phenotype. This phenotype had been so far putatively invisible to the current detection methods based on fluorescent microscopy, whereas negativity of those cells for CD45, CD31 and α-SMA as well as size and morphology allow to classify them as potential CTCs. The putative K-V-CTCs were indeed the size of epithelial CTCs, revealed more frequently protrusions and correlated with the higher number of involved lymph nodes. Further studies should be performed to confirm cancer-origin of those cells, understand their biology, dissect markers allowing their unquestionable detection and validate their clinical relevance.

Interestingly, two other EMT-related types of CTCs, i.e. epithelial-mesenchymal and mesenchymal, seemed to be smaller than epithelial CTCs, which should be taken into account for size-based methods of CTCs detection and investigated carefully in regard to inclusion criteria for CTCs classification. In total, EMT-related CTCs appeared to correlate with some aspects of tumor progression, among others progression of tumors under the systemic treatment. As EMT signatures were proven to coexist with stemness program, it could potentially explain resistance of those particular clones of tumor cells to therapy ^26^.

In the current study we observed various patterns of tumor dissemination. Single and clustered CTCs were found, as well as CTCs clustering with normal cells of undefined type and origin. Although those clusters were identified in only 8% of patients, they were associated with the greater number of metastatic sites when compared to CTCs-negative patients and patients with single CTCs. It corroborates data showing that CTCs clusters alone and together with normal cells e.g. neutrophils possess higher capacity to form metastases ^27, 28^. Approximately 40% of CTCs-positive patient samples were characterized by heterotypic seeding, with more than one phenotype of CTCs detected. Epithelial phenotype occurred the most frequently in TNBC and in cases with multiple coexisting bone and visceral metastases. EMT-related CTCs were more frequently detected in LumB/Her2+ BC subtype and specifically mesenchymal CTCs were found exclusively in cases with visceral metastases. It might be hypothesized that mechanism of tumor dissemination is different among molecular subtypes, e.g. luminal BC might be more prone to EMT. Similarly, different secondary organs might attract or seed CTCs of different phenotype. However, all those observations need to be verified in larger study cohorts.

cCAFs, characterized as α-SMA+/K-/V- or V+/CD45&CD31-cells were co-detected with CTCs, especially those of epithelial and negative phenotype, in 7 (3%) BC, mainly M1, patients. In contrast, using the same α-SMA as a marker of identification, cCAFs were previously identified by Ao et al. in 88.2% of metastatic BC patients and 23.1% of local BC patients, whereas Ortiz-Otero et all. detected cCAFs in all metastatic BC patients ^6, 29^. They were also found at the higher numbers per patient than in our study. One explanation for such a difference in cCAFs yield might be the study cohort and methodology as during our study we enrolled only 26 M1 patients and in our protocol cells were stained in suspension which might generate hypothetically greater loss of cells in contrast to staining performed on microfilter ^6^ or glass microscope slides performed by the others ^29^. Interestingly, the majority of cCAFs detected in our study were negative for vimentin. In line with this observation, some studies showed that loss of vimentin enhances fibroblasts’ motility through microchannels ^30^, reduced number and size of cell–matrix adhesions ^31^ and cell stiffness ^32^, which may potentially allow them to enter blood stream. Interestingly, no CD29+ cCAFs were found, whereas this subtype of cCAFs was found predominantly within primary tumors of luminal BC ^33^. Due to the small number of cCAFs-positive patients, statistics performed on those samples lacked statistical power. Nevertheless, it was noted that the frequency of metastases was greater in patients with coexisting CTCs and cCAFs in comparison to patients with no CTCs or only CTCs. cCAFs were also present in patients with exclusively visceral metastases, while, to date, there is no study connecting cCAFs presence with the type of metastases.

Last but not least, it is important to note that to date only a few studies have assessed the suitability of the imFC system for CTC detection ^34–38^ and only one has been applied to BC patients, however, at the low number of cases (n=8) ^39^. Thus, our study is the first using imFC on the relatively large population of patients and focused on multiparameter analysis of those rare cells. Of note, imFC with its improved - in comparison to CellSearch - resolution allowed not only to define CTCs but also examine their morphological details, which in the future might even upgrade the criteria of CTC identification ^40^.

## Conclusions

To sum up, to the best of our knowledge this is the first study showing the feasibility of imFC to co-detect CTCs and cCAFs in peripheral blood collected from BC patients in one-tube assay. Both CTCs of different phenotype and cCAFs might indicate more advanced disease, which merits further investigation in larger cohorts of patients.

## Data Availability

The data that support the findings of this study are available from the corresponding author upon reasonable request.

## Supplementary materials

**Supplemental Table 1:** Antibodies dilution and acquisition parameters used for ImFC.

**Supplemental Table 2:** Correlations of CTC phenotypes with cCAFs.

**Supplemental Table 3**: Comparison of exclusive CTC phenotypes among clinico-pathological features and response to treatment of patients with breast cancer

**Supplemental Table 4:** Distribution of dominant CTC phenotypes among clinico-pathological features and response to treatment of patients with breast cancer.

**Fig. S1:** Representative pictures of different breast cancer (BC) cell lines corresponding different BC molecular subtypes envisioned by imaging flow cytometry.

## Authors contribution

Conceptualization, A.J.Z., A.Ma. and N.B.-K.; methodology, A.Mu. and N.B.-K.; formal analysis, A.Mu., R.W., W.Ś. and N.B.-K.; investigation, A.Mu., R.W., W.Ś. and N.B.-K.; resources, E.S., and A.J.Z.; data curation, A.Mu., R.W., and G.S.; writing—original draft preparation, A.Mu. and N.B.-K.; writing—review and editing, all authors; visualization, A.Mu. and N.B.-K.; supervision, A.J.Z. and N.B.-K.; project administration, A.J.Z.; funding acquisition, A.J.Z., A.Ma. and N.B.-K.

## Research Funding

This research was funded by National Center for Research and Development, Poland (grant number WPC/33/HESCAP/2018 for A.Z.). The funding organizations played no role in the design of study, choice of enrolled patients, review and interpretation of data, preparation of manuscript, or final approval of manuscript.

## Acknowledgements

We thank all patients who agreed to donate blood, Dr Beata Pieczyńska-Uziębło for technical assistance to stain samples, Barbara Bollin-Matysiak for technical assistance with CTC isolation and clinical data curation as well as Justyna Topa for technical assistance with CTC isolation. Further, we would like also to acknowledge Peter Grešner for providing statistics consultation within the services of the Centre of Biostatistics and Bioinformatics Analysis at the Medical University of Gdańsk, Poland, working as part of “Excellence Initiative — Research University” (grant no. MNiSW 07/IDUB/2019/94).

