## Supplementary material for "Improved characterization of circulating tumor cells and cancer-associated fibroblasts in breast cancer patients using imaging flow cytometry": Fig. S1

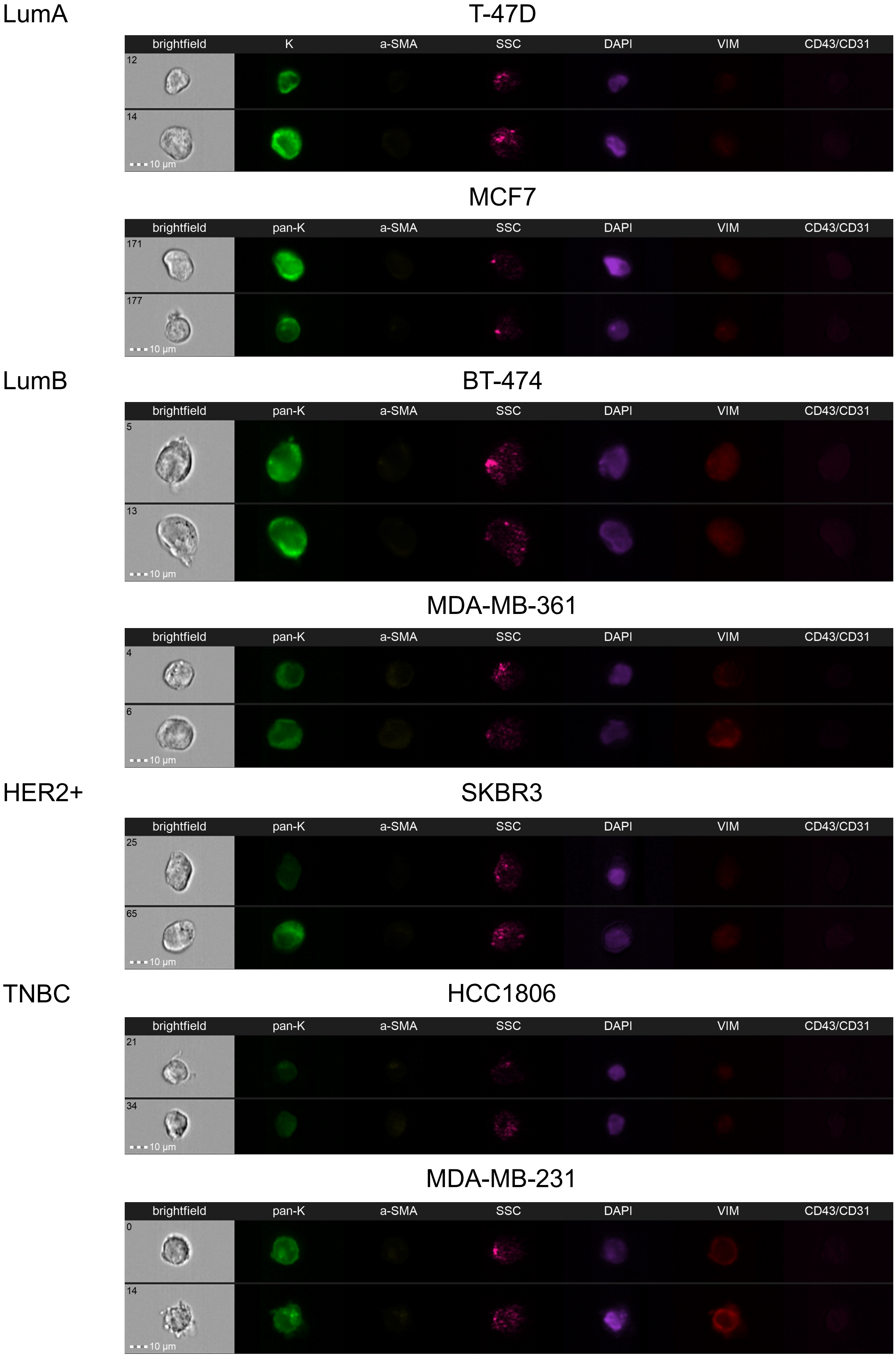


**Fig. S1** Representative pictures of different breast cancer (BC) cell lines corresponding different BC molecular subtypes envisioned by imaging flow cytometry.
