## Supplemental Table 1 for "Improved characterization of circulating tumor cells and cancer-associated fibroblasts in breast cancer patients using imaging flow cytometry"

Antibodies dilution and acquisition parameters used for ImFC.

| **Antibody** | **Company** | **Cat. nr.** | **Fluorophor** | **Dilution** | **Laser** | **Laser power** | **Channel** |
| --- | --- | --- | --- | --- | --- | --- | --- |
| BF |  |  |  |  |  |  | 1+9 |
| pan-keratin (AE1/AE3) | Thermo Fisher Scientific | 53-9003-82 | Alexa 488 | 1:2500 | 488 | 100mW | 2 |
| Pan-keratin (C11) | Thermo Fisher Scientific | MA5-18156 | Alexa 488 | 1:2500 | 488 | 100mW | 2 |
| a-SMA (1A4) | R&Dsystems | IC1420P | PE | 1:50 | 488 | 100mW | 3 |
| SSC |  |  |  |  | 785 |  | 6 |
| DAPI | BD Biosciences | 564907 | DAPI | 0,04ug/ml | 405 | 100mW | 7 |
| CD29 (TS2/16) | Thermo Fisher Scientific | 63-0299-42 | SuperBright600 | 1:1000 | 405 | 100mW | 10 |
| Vimentin (D21H3) | Cell Signaling | 9856 | Alexa 647 | 1:100 | 647 | 150mW | 11 |
| CD45 (REA747) | Miltenyi Biotec | 130-110-635 | APC-Vio770 | 1:50 | 647 | 150mW | 12 |
| CD31 (WM59) | BioLegend | 303120 | APC-Cy7 | 1:10 | 647 | 150mW | 12 |
