## Supplemental Table 2 for "Improved characterization of circulating tumor cells and cancer-associated fibroblasts in breast cancer patients using imaging flow cytometry"

Correlations of CTC phenotypes with cCAFs.

|  |  | **epi CTCs** | **epi-mes CTCs** | **mes CTCs** | **neg CTCs** | **cCAFs** |
| --- | --- | --- | --- | --- | --- | --- |
| **epi CTCs** | R^2^ | 1 | -0.015 | **0.259** | **0.772** | **0.165** |
|  | p-values |  | 0.813 | **<0.001** | **<0.001** | **0.017** |
|  | n | 210 | 210 | **210** | **210** | **210** |
| **epi-mes CTCs** | R^2^ |  | 1 | 0.086 | 0.105 | 0.086 |
|  | p-values |  |  | 0.215 | 0.128 | 0.214 |
|  | n |  | 210 | 210 | 210 | 210 |
| **mes CTCs** | R^2^ |  |  | 1 | **0.314** | 0.097 |
|  | p-values |  |  |  | **<0.001** | 0.163 |
|  | n |  |  | 210 | **210** | 210 |
| **neg CTCs** | R^2^ |  |  |  | 1 | **0.250** |
|  | p-values |  |  |  |  | **<0.001** |
|  | n |  |  |  | 210 | **210** |
| **cCAFs** | R^2^ |  |  |  |  | 1 |
|  | p-values |  |  |  |  |  |
|  | n |  |  |  |  | 210 |
