## Supplemental Table 3-4 for "Improved characterization of circulating tumor cells and cancer-associated fibroblasts in breast cancer patients using imaging flow cytometry"

Comparison of exclusive CTC phenotypes among clinico-pathological features and response to treatment of patients with breast cancer. Chi squared test was performed. Due to missing data not all numbers sum up to 210.

|  |  | **Exclusive CTC phenotypes** | | | | |  |
| --- | --- | --- | --- | --- | --- | --- | --- |
| Variable | total n | epi | mes | epi-mes | neg | heterogenous | p-value |
| **Age** |  |  |  |  |  |  |  |
| <50 | 16 | 2 (18.2%) | 2 (12.5%) | 2 (66.75) | 0 (0%) | 10 (43.5%) | **0.02** |
| >>50 | 42 | 9 (81.8%) | 14 (87.5%) | 1 (33.3%) | 5 (100%) | 13 (56.5%) |  |
| **cT stage** |  |  |  |  |  |  |  |
| cT1-2 | 27 | 5 (62.5%) | 8 (72.7%) | 2 (100%) | 2 (50%) | 10 (66.7%) | 0.8 |
| cT3-4 | 13 | 3 (37.5%) | 3 (27.3%) | 0 (0%) | 2 (50%) | 5 (33.3%) |  |
| **cN stage** |  |  |  |  |  |  |  |
| 0 | 15 | 5 (62.5%) | 4 (40%) | 2 (100%) | 0 (0%) | 4 (25%) | 0.11 |
| 1 | 25 | 3 (37.5%) | 6 (60%) | 0 (0%) | 4 (100%) | 12 (75%) |  |
| **M stage** |  |  |  |  |  |  |  |
| 0 | 46 | 7 (63.6%) | 12 (75%) | 3 (100%) | 5 (100%) | 19 (82.6%) | 0.05 |
| 1 | 12 | 4 (36.4%) | 4 (25%) | 0 (0%) | 0 (0%) | 4 (15.4%) |  |
| **Grading** |  |  |  |  |  |  |  |
| 1 | 3 | 1 (11.1%) | 1 (8.3%) | 0 (0%) | 0 (0%) | 1 (5%) | 0.46 |
| 2 | 26 | 3 (33.3%) | 7 (58.3%) | 3 (100%) | 4 (100%) | 9 (45 %) |  |
| 3 | 19 | 5 (55.6%) | 4 (33.3%) | 0 (0%) | 0 (0%) | 10 (50%) |  |
| **Molecular type** |  |  |  |  |  |  |  |
| Luminal A | 9 | 1 (10%) | 4 (25%) | 1 (33.3%) | 1 (25%) | 2 (8.7%) | 0.1 |
| Luminal B HER2- | 14 | 1 (10%) | 4 (25%) | 2 (66.7%) | 0 (0%) | 7 (30.4%) |  |
| Luminal B HER2+ | 13 | 2 (20%) | 4 (25%) | 0 (0%) | 3 (75%) | 4 (17.4%) |  |
| Non luminal HER2+ | 3 | 0 (0%) | 1 (6.3%) | 0 (0%) | 0 (0%) | 2 (8.7%) |  |
| Triple Negative | 17 | 6 (60%) | 3 (18.8%) | 0 (0%) | 0 (0%) | 8 (34.8%) |  |
| **Response to treatment** |  |  |  |  |  |  |  |
| stable disease | 6 | 0 (0%) | 2 (25%) | 1 (100%) | 2 (50%) | 1 (8.3%) | 0.45 |
| complete response | 9 | 2 (50%) | 1 (12.5%) | 0 (0%) | 1 (25%) | 5 (41.7%) |  |
| partial response | 13 | 2 (50%) | 5 (62.5%) | 0 (0%) | 1 (25%) | 5 (41.7%) |  |
| progressive disease | 1 | 0 (0%) | 0 (0%) | 0 (0%) | 0 (0%) | 1 (8.3%) |  |

**Supplemental Table 4**

Distribution of dominant CTC phenotypes among clinico-pathological features and response to treatment of patients with breast cancer. Chi squared test was performed. Due to missing data not all numbers sum up to 210.

|  |  | **Dominant CTC phenotypes** | | | | | |  |
| --- | --- | --- | --- | --- | --- | --- | --- | --- |
| Variable | total n | epi | mes | epi-mes | neg | heterogenous | | p-value |
| **Age** |  |  |  |  |  | |  |  |
| <50 | 16 | 6 (27.3%) | 3 (17.6%) | 4 (66.7%) | 2 (20%) | | 1 (33.3%) | 0.08 |
| >>50 | 42 | 16 (73.7%) | 14 (82.4%) | 2 (33.3%) | 8 (80%) | | 2 (66.7%) |  |
| **cT stage** |  |  |  |  |  | |  |  |
| cT1-2 | 27 | 10 (71.4%) | 8 (66.7%) | 3 (100%) | 4 (50%) | | 2 (66.7%) | 0.65 |
| cT3-4 | 13 | 4 (28.6%) | 4 (33.3%) | 0 (0%) | 4 (50%) | | 1 (33.3%) |  |
| **cN stage** |  |  |  |  |  | |  |  |
| 0 | 15 | 6 (40%) | 4 (36.4%) | 2 (66.7%) | 2 (25%) | | 1 (33.3%) | 0.88 |
| 1 | 25 | 9 (60%) | 7 (63.6%) | 1 (33,3%) | 6 (75%) | | 2 (66.7%) |  |
| **M stage** |  |  |  |  |  | |  |  |
| 0 | 46 | 16 (72.7%) | 13 (76.5%) | 4 (66.7%) | 10 (100%) | | 3 (100%) | **0.03** |
| 1 | 12 | 6 (23.1%) | 4 (23.5%) | 2 (33.3%) | 0 (0%) | | 0 (0%) |  |
| **Grading** |  |  |  |  |  | |  |  |
| 1 | 3 | 2 (11.1%) | 1 (7.7%) | 0 (0%) | 0 (0%) | | 0 (0%) | 0.84 |
| 2 | 26 | 8 (44.4%) | 7 (53.8%) | 4 (80%) | 6 (66.7%) | | 1 (33.3%) |  |
| 3 | 19 | 8 (44.4%) | 5 (38.5%) | 1 (20%) | 3 (33.3%) | | 2 (66.7%) |  |
| **Molecular type** |  |  |  |  |  | |  |  |
| Luminal A | 9 | 2 (9.5%) | 4 (23.5%) | 1 (16.7%) | 2 (22.2%) | | 0 (0%) | 0.17 |
| Luminal B HER2- | 14 | 4 (19%) | 4 (23.5%) | 3 (50%) | 1 (11.1%) | | 2 (66.7%) |  |
| Luminal B HER2+ | 13 | 3 (14.3%) | 4 (23.5%) | 2 (33.3%) | 4 (44.4%) | | 0 (0%) |  |
| Non luminal HER2+ | 3 | 1 (4.8%) | 1 (5.9%) | 0 (0%) | 1 (11.1%) | | 0 (0%) |  |
| Triple Negative | 17 | 11 (52.4%) | 4 (23.5%) | 0 (0%) | 1 (11.1%) | | 1 (33.3%) |  |
| **Response to treatment** |  |  |  |  |  | |  |  |
| stable disease | 6 | 0 (0%) | 2 (25%) | 2 (100%) | 2 (28.6%) | | 0 (0%) | 0.19 |
| complete response | 9 | 4 (44.4%) | 1 (12.5%) | 0 (0%) | 3 (42.9%) | | 1 (33.3%) |  |
| partial response | 13 | 4 (44.4%) | 5 (62.5%) | 0 (0%) | 2 (28.6%) | | 2 (66.7%) |  |
| progressive disease | 1 | 1 (11.1%) | 0 (0%) | 0 (0%) | 0 (0%) | | 0 (0%) |  |
